# Errors of Interpretation - ‘*Correcting the record on the comparative efficacy of surgical masks versus respirators: Historical research findings suggesting their equivalence and used to support downgraded respiratory protection for non-ICU UK healthcare workers, resulted from unrecognised errors of arithmetic*’

**DOI:** 10.1101/2022.07.15.22277662

**Authors:** Martyn AC Wilkinson, Evonne T Curran, David R Tomlinson

## Abstract

The first UK ‘COVID-19 Guidance for Infection Prevention and Control (IPC) in healthcare settings’, March 2020, restricted respiratory protection equipment (RPE) to aerosol generating procedures (AGPs) and the ICU. Supportive data in a linked meta-analysis stated equivalent protection – i.e., 80% - afforded by fluid resistant surgical face masks (FRSMs) and RPE, against SARS-CoV-1 infection.

Visual inspection of the meta-analysis data suggested greater healthcare worker (HCW) protection from RPE. Detailed statistical analysis confirmed significantly greater HCW protection from RPE (p<0.0009) compared to FRSMs: sadly, the meta-analysis had been subject to calculation error.

Research data supporting the restriction of RPE to AGPs and the ICU were flawed. Consequently, non-ICU HCWs have been inappropriately exposed to SARS-CoV-2 aerosol inhalation transmission.

To minimise HCW occupational COVID-19, UK IPC guidance should be changed to require RPE during all confirmed / suspected infectious SARS-CoV-2 patient care.

## Introduction

The March 2020 UK COVID-19 Guidance for infection prevention and control in healthcare settings ‘Version 1’^1^ contained three foundational errors which have been perpetuated in every subsequent iteration. The first error was that SARS-CoV-2 transmission was considered to occur predominantly via large respiratory droplets: this statement has no suitable evidence base. The second error was that inhalation transmission risk was restricted to a short list of so-called aerosol generating procedures (AGPs)^1^. However, this statement is contra to decades of research into human bioaerosol production confirming their abundant release during activities such as coughing, sneezing and speech, with respiratory viral genomic material even identified in exhaled breath samples from infectious adults and children^2^. Recently, AGPs have been proven to generate far fewer aerosols than normal respiratory activities^3-4^: for SARS-CoV-2, the dominant mechanism for transmission is via aerosol inhalation^5^.

The third error, and the topic of this letter, was the suggestion that fluid resistant surgical face masks (FRSMs) conferred equivalent proportionate risk of infection reduction to the wearer as respiratory protective equipment (RPE; FFP3 / N95)^1^. Indeed, the COVID-19 guidance stated that the use of both surgical face masks and RPE were associated with ∼80% reduction in risk^1^. This statement was attributed to a meta-analysis by Offeddu et al (2017)^6^, in which 2 tables and 2 forest plots provided supporting evidence for this specific claim.

On face value, this conclusion is problematic: it is against physical laws concerning bioaerosol filtration, and laboratory data demonstrate significantly greater filtration performance and protection afforded by RPE^7^. On closer inspection, it appears the authors made an error in their calculation of the odds ratios [Figure 5 B; 5C]^6^: the weighted odds ratio of 0.12 for N95 vs no mask looks suitable, but the odds ratio of 0.13 for medical masks vs no mask is incorrect – it should be 0.29; presumably an error in the execution of this calculation. In view of these concerns, we reanalysed data presented by Offeddu et al^6^.

## Methods

As data were gathered from different sources and combined, a non-parametric statistical test was chosen to analyse the difference in the proportion of HCWs infected with SARS when wearing no mask, medical masks, or N95 respirators. The Infer package in R (R version 4.2.0)^8^ was used to perform permutation tests on the difference in proportions infected, and to construct bootstrap 95% confidence intervals for the difference in proportions. The sample size employed for each permutation test and bootstrap procedure was 10000. When no permutations from the null distribution were as, or more, extreme than the measured value, the upper bound for the p value was taken as the upper 95% bound for the probability of success when no successes were measured in 10000 Bernoulli trials; otherwise, the p value was taken as the proportion of permuted samples in the null distribution that were as, or more, extreme than the measured value. Holm’s correction was applied to the p values to control the family-wise error rate due to multiple comparisons.

## Results

The key results are presented in Table 1, with additional analyses here [supplement]. The difference in the proportions experiencing SARS infection following the use of medical mask versus no mask during SARS patient care – 8.0% - just achieved statistical significance (0.049), in keeping with a small protective effect from medical mask use. However, the difference in the proportions infected comparing N95 mask use versus no masks - 31% - was highly significant (p <0.0009), in favour of N95 use. Consequently, the difference in the proportion infected comparing N95 mask use versus medical masks – 12% - was also highly significant (p<0.0009), demonstrating optimal healthcare worker protection from N95 mask use. This finding that N95 masks provided superior protection against infection is particularly notable, since the baseline risk in the non-intervention group was greater in studies involving N95, compared to medical mask use (36.8% vs 25.1% of unmasked infected, respectively).

**Table 1.** Comparison in the difference between different masks and the proportion infected.

| Comparison | Difference in proportion infected | 95% CI | P value |
| --- | --- | --- | --- |
| None vs Medical Mask | 0.0796 | (0.0038, 0.158) | 0.049 |
| None vs N95 | 0.312 | (0.240, 0.385) | < 0.0009 |
| Medical mask vs N95 | 0.117 | (0.0618, 0.17) | < 0.0009 |

## Discussion

An apparent error of arithmetic led us to reanalyse a 2017 metanalysis^6^ describing the comparative protective effect of medical masks versus N95 respirators. We demonstrate that, while a medical mask clearly affords some protection to the wearer, there was a significantly greater protective effect during N95 mask use. These data are clearly in keeping with physical laws concerning airborne particle filtration, and historical laboratory data proving significantly greater efficacy of RPE compared to medical masks. Furthermore, these findings are also in line with any technology better able to mitigate the dominant mechanism for SARS-CoV-2 transmission – i.e., aerosol inhalation^5^.

This first and every subsequent UK COVID-19 IPC guidance employing Offeddu et al^6^ was importantly flawed and therefore unsafe. We believe these findings should prompt an urgent revision to any infection prevention and control guidance drawing upon Offeddu et al^6^; in short, all SARS-CoV-2 facing healthcare staff require RPE for optimal protection in the workplace.

Sadly, there are other flaws underlying this first UK guidance which remain uncorrected. Most importantly, although authors state that guidance is based on “the reasonable assumption that the transmission characteristics of COVID-19 are similar to those of the 2003 SARS-CoV outbreak”, there is no acknowledgement that instances of aerosol transmission outwith AGPs were a proven component of human-to-human SARS Coronavirus transmission^9^. Also, guidance^10^ continues to ignore the aerosol creating potential of normal respiratory behaviours such as breathing and coughing, and their associated transmission risk.

### Methodological note

Our analyses differed to those of Offeddu *et al*^6^ in one additional respect: we used pooled data, without weighting. However, this will have minimal impact on the results given the magnitude of superiority of N95 masks over medical masks described. We have also carefully re-checked our calculations and confirm that we can find no errors.

## Conclusion

The first UK COVID-19 IPC guidance^1^ and all subsequent editions contain 3 foundational errors on transmission, undermining HCW safety:

1. There was no data to support the suggestion that SARS-CoV-2 transmission was predominantly via large droplets. However, robust historical data on SARS Coronavirus confirmed its airborne nature out-with AGPs.
2. The term ‘AGP’ is scientifically invalid, given that normal respiratory activities generate orders of magnitude greater aerosols than intubation – the AGP considered to be of greatest transmission risk. Accordingly, any IPC guidance restricting RPE use to AGPs is dangerously flawed.
3. RPE affords significantly greater protection to the wearer, not, as stated, equivalence with FRSMs.

We urge IPC guidance authors to acknowledge and respond to these errors and provide updated guidance recommending RPE for all HCWs during known / suspected infectious SARS-CoV-2 patient care.

## Data Availability

All data produced in the present work are contained in the manuscript

