## supplement for "Errors of Interpretation - ‘*Correcting the record on the comparative efficacy of surgical masks versus respirators: Historical research findings suggesting their equivalence and used to support downgraded respiratory protection for non-ICU UK healthcare workers, resulted from unrecognised errors of"

Method

Because data were gathered from different sources and combined, a non-parametric statistical test was chosen to analyse the difference in the proportion of HCWs infected with SARS when wearing no protection, medical masks, or N95 respirators. The Infer package in R (R version 4.2.0) was used to perform permutation tests on the difference in proportions infected, and to construct bootstrap 95% confidence intervals for the difference in proportions. The sample size employed for each permutation test and bootstrap procedure was 10000. When no permutations from the null distribution were as, or more, extreme than the measured value, the upper bound for the p value was taken as the upper 95% bound for the probability of success when no successes were measured in 10000 Bernoulli trials; otherwise, the p value was taken as the proportion of permuted samples in the null distribution that were as, or more, extreme than the measured value. Holm's correction was applied to the p values to control the family-wise error rate due to multiple comparisons.

Results

Medical masks vs no mask

Table S1 shows the data used for comparison between HCWs using medical masks and those using no protection.

Table S1

| Mask type | Number infected (total) | Proportion infected |
|---|---:|---:|
| Medical mask | 56 (326) | 0.172 |
| None | 45 (179) | 0.251 |

The difference in proportion infected was 0.0796 (95% CI: 0.0038, 0.158), $p = 0.049$. Figure S1 displays the permuted null distribution of the difference in proportion together with the actual measured difference (red line); the proportion of the null distribution with values as (or more) extreme than the measured difference are shaded pink. Figure S2 depicts the bootstrap samples of the actual difference in proportion; the green lines are the 95% confidence limits, and the 95% confidence interval is shaded green.

Figure S1

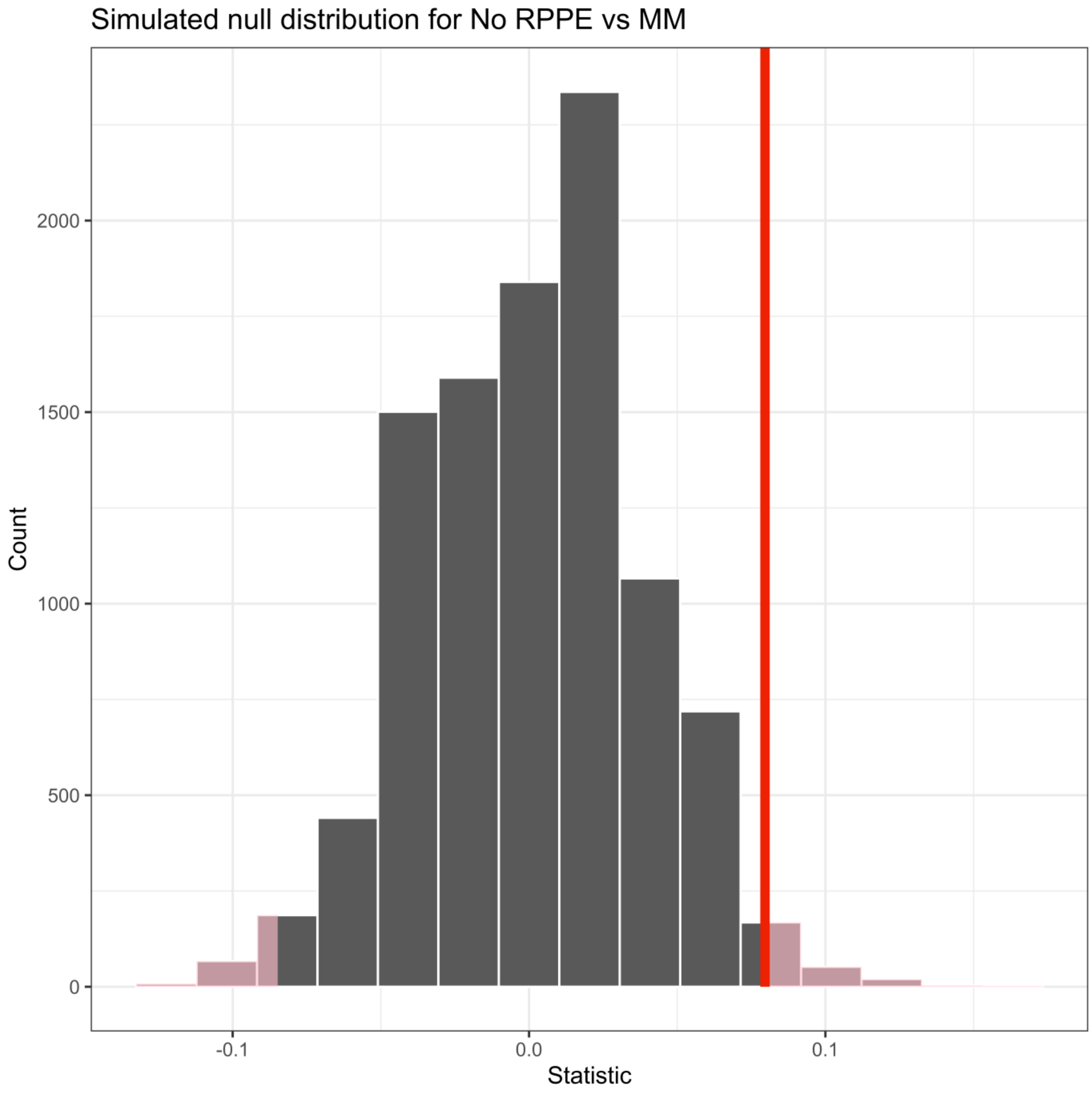

Figure S2

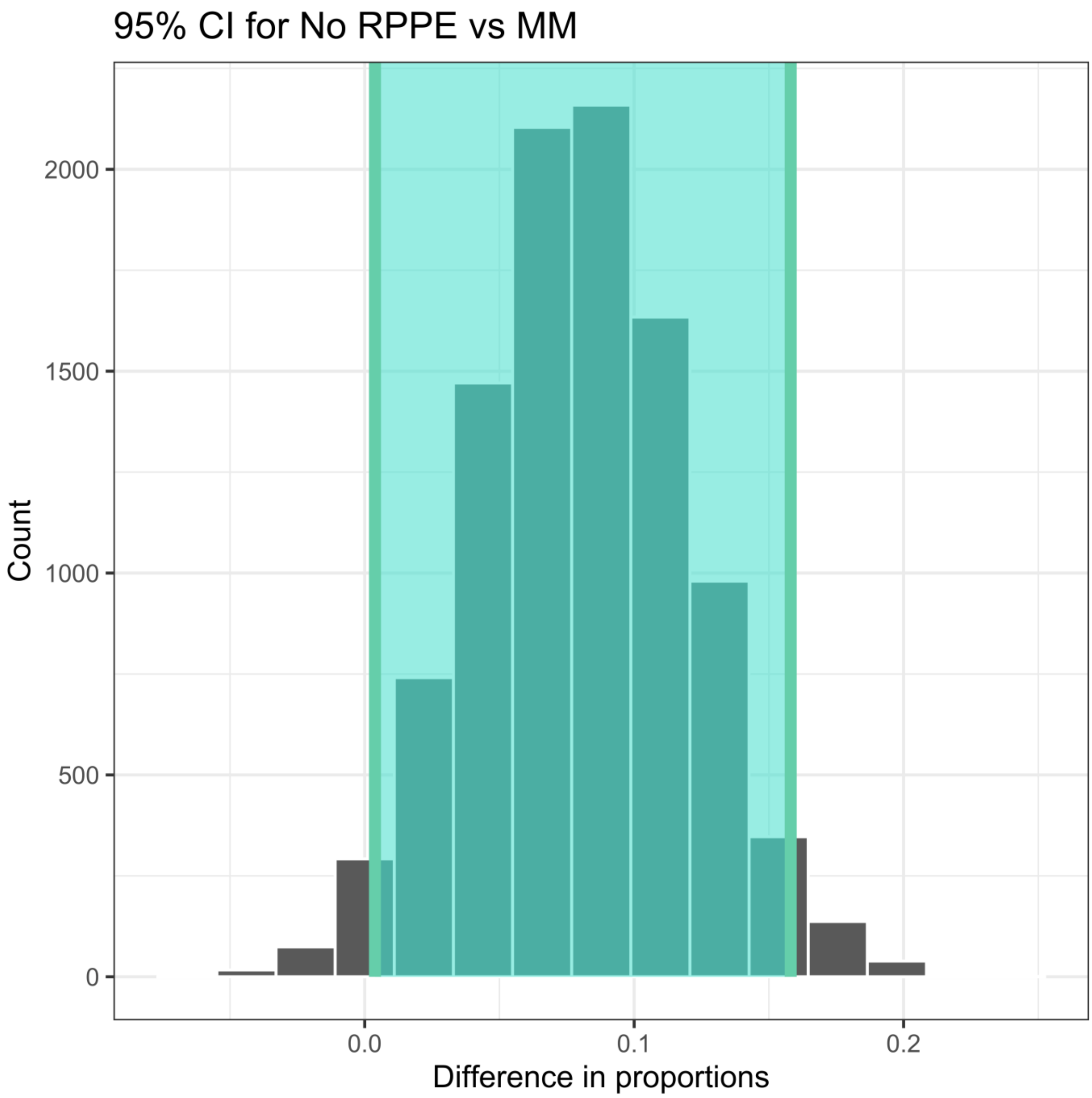

95% CI for No RPPE vs MM
Count
2000
1500
1000
500
0
0.0
0.1
0.2
Difference in proportions

N95 vs no mask

Table S2 shows the data used for comparison between HCWs using N95 respirators and those using no protection.

Table S2

| Mask type | Number infected (total) | Proportion infected |
|---|---:|---:|
| N95 | 9 (163) | 0.0552 |
| None | 86 (234) | 0.368 |

The difference in proportion infected was 0.312 (95% CI: 0.240, 0.385), $p < 0.0009$. Figure S3 displays the permuted null distribution of the difference in proportion, while Figure S4 depicts the bootstrap samples of the actual difference in proportion along with 95% confidence interval.

Figure S3

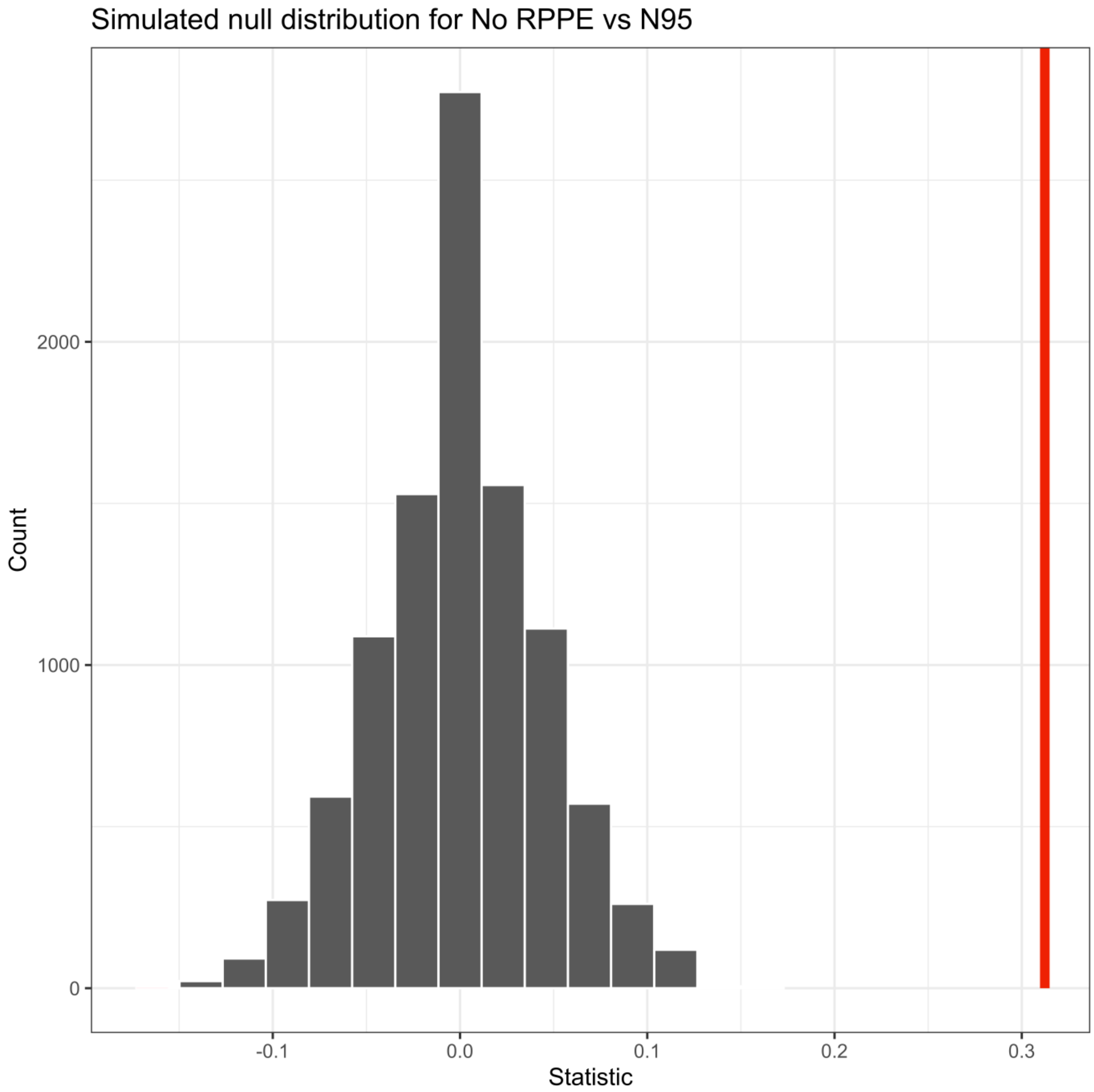

Simulated null distribution for No RPPE vs N95
Count
2000
1000
0
-0.1
0.0
0.1
0.2
0.3
Statistic

Figure S4

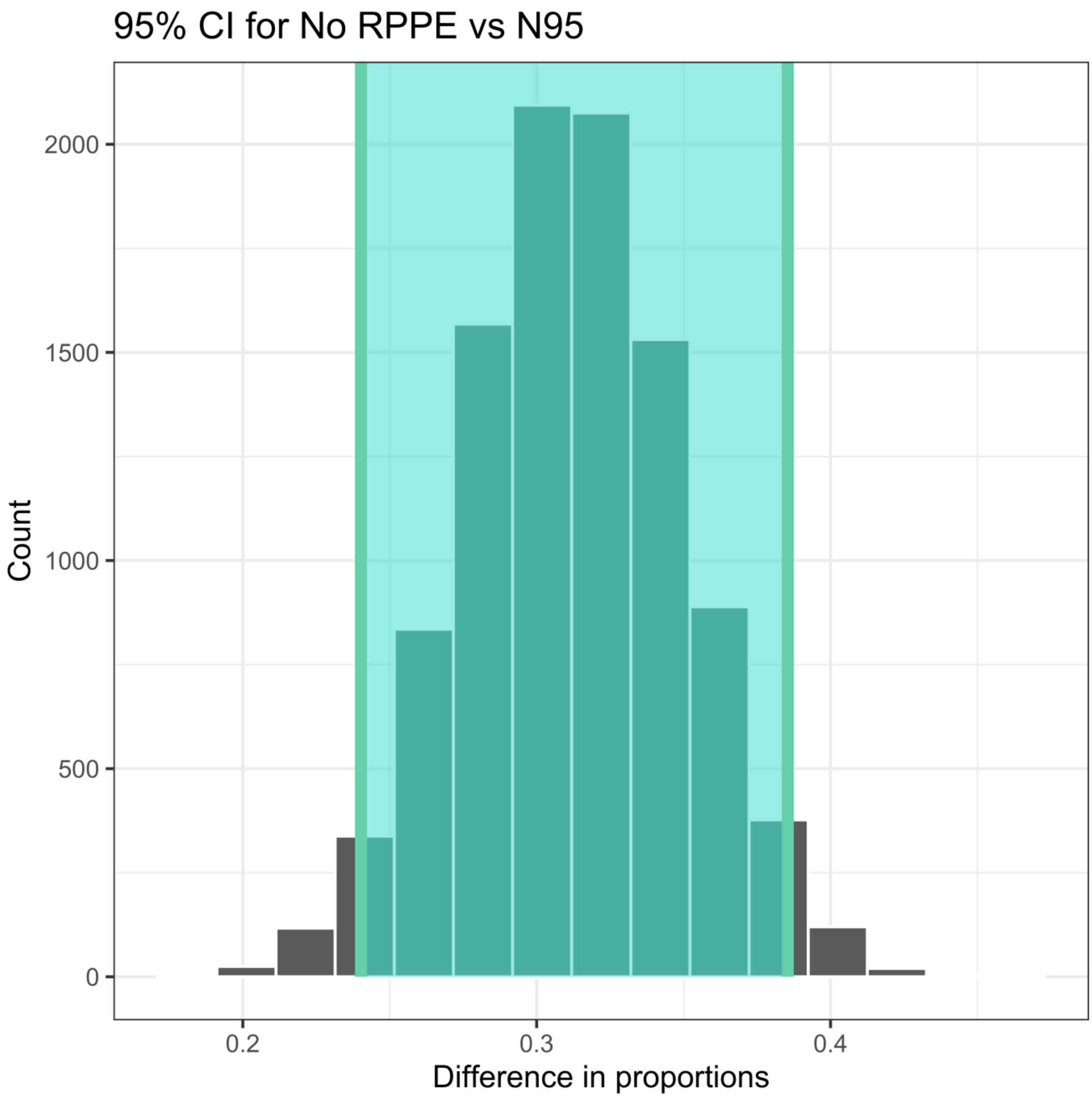

95% CI for No RPPE vs N95
2000
1500
1000
500
0
Count
0.2
0.3
0.4
Difference in proportions

Medical mask vs N95

Table S3 shows the data used for comparison between HCWs using N95 respirators and those using medical masks.

Table S3

| Mask type | Number infected (total) | Proportion infected |
|---|---|---|
| Medical mask | 56 (326) | 0.172 |
| N95 | 9 (163) | 0.0552 |

The difference in proportion infected was 0.117 (95% CI: 0.0618, 0.170), $p < 0.0009$. Figure S5 displays the permuted null distribution of the difference in proportion, while Figure S6 depicts the bootstrap samples of the actual difference in proportion along with 95% confidence interval.

Figure S5

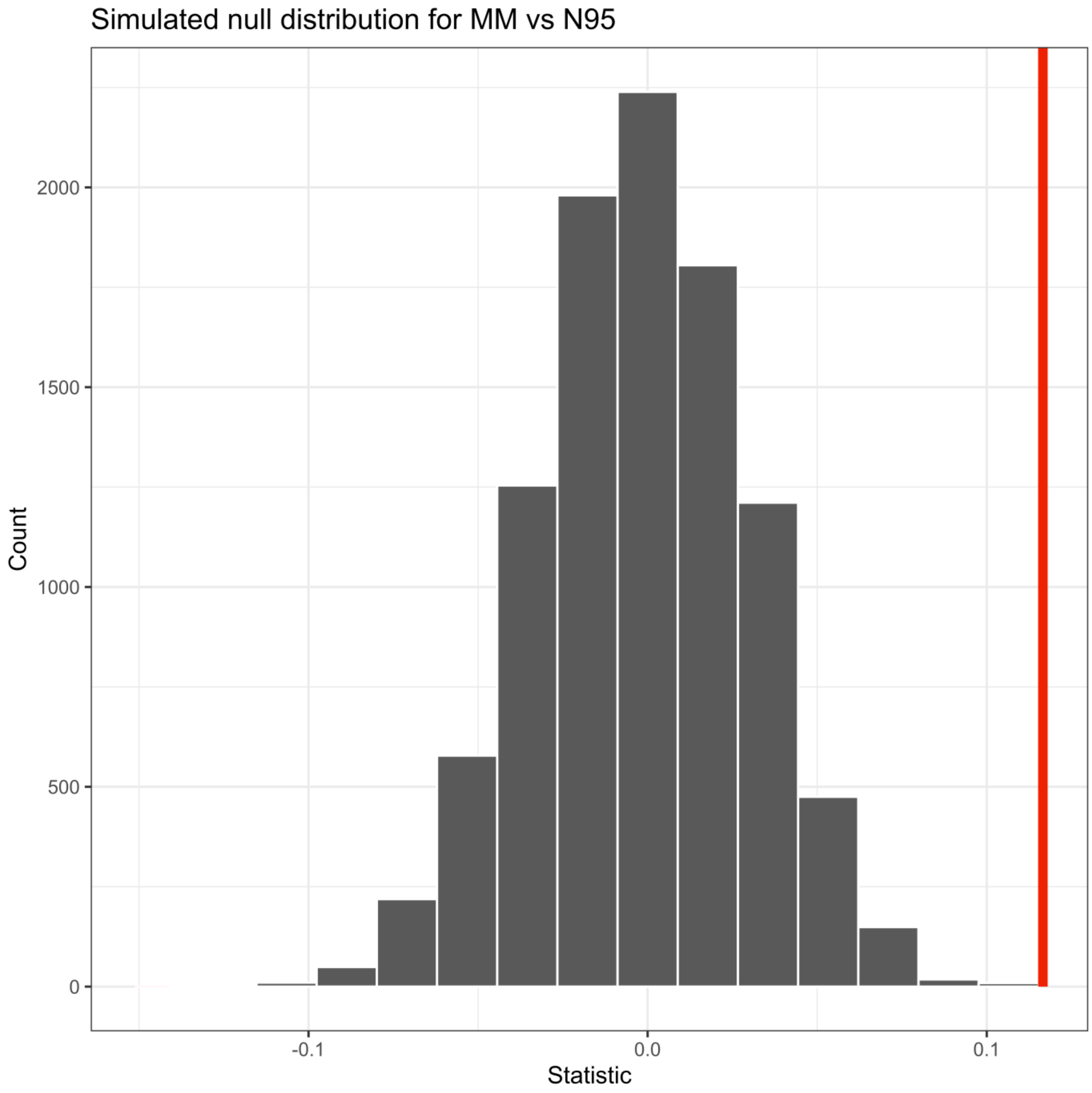

Simulated null distribution for MM vs N95
Count
2000
1500
1000
500
0
-0.1
0.0
0.1
Statistic

Figure S6

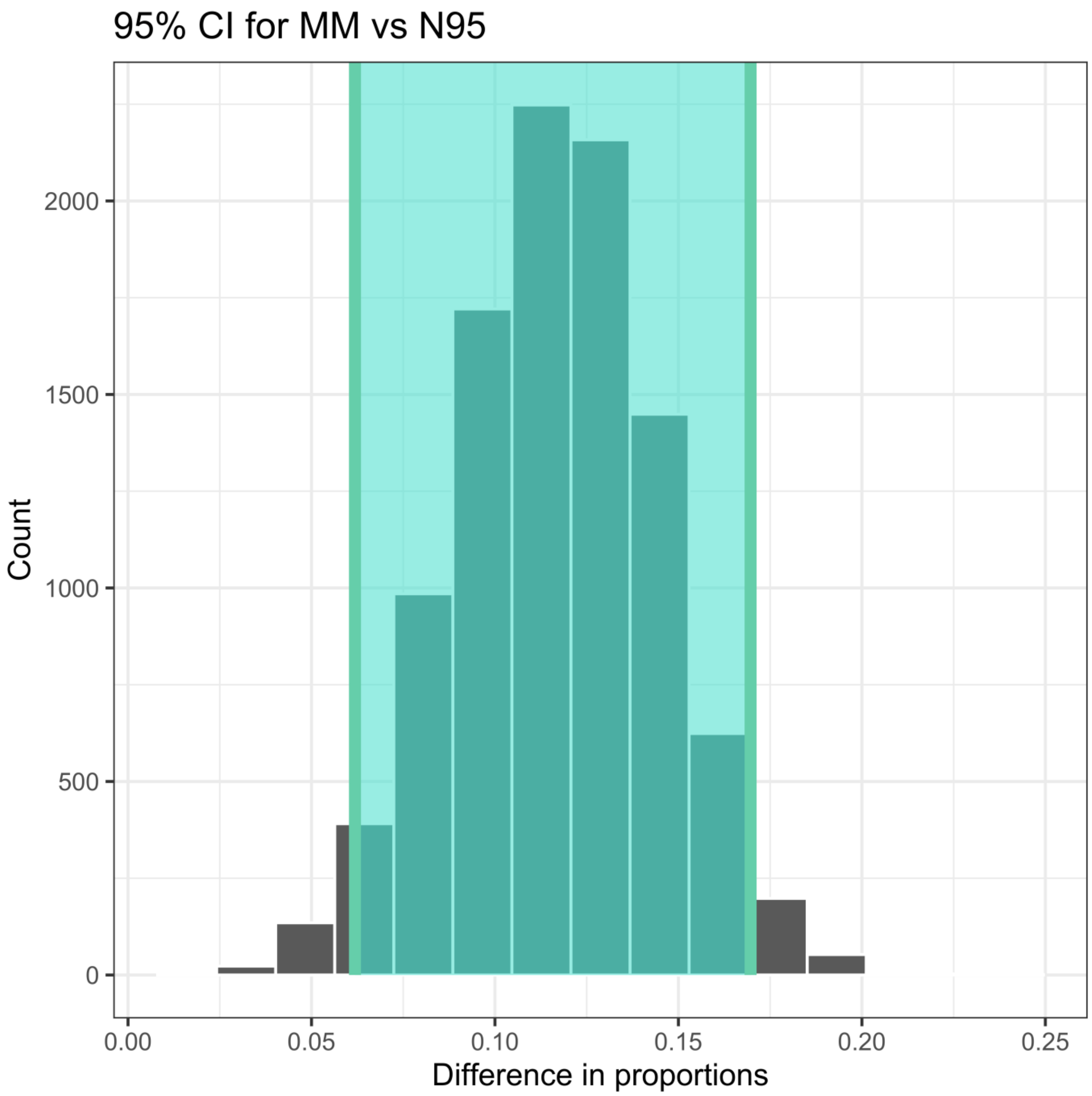
95% CI for MM vs N95
Count
2000
1500
1000
500
0
0.00
0.05
0.10
0.15
0.20
0.25
Difference in proportions

Discussion

When comparing the protection of HCWs against infection with SARS, medical masks offered a 7.9 percentage point reduction in the percentage infected, compared to no masks; the percentage reduction was 31.5%. For N95 respirators, the percentage point reduction was 31.2 in the percentage infected, compared to no masks; the percentage reduction was 85%. N95 respirators gave an 11.7 percentage point reduction in the percentage infected, compared to medical masks; the percentage reduction was 67.9%. It is notable that the proportion infected wearing no protection was much higher during the comparison with N95 respirators than during the comparison with medical masks; assuming that there was little community transmission during the SARS outbreak, it seems likely that the HCWs were in a more risky clinical environment during the N95 comparison period.
Table S4 displays a summary of the findings for the comparison between mask types regarding protection against SARS infection.

Table S4

| Comparison | Difference in proportion | 95% CI | P value |
|---|---:|---:|---:|
| None vs Medical mask | 0.0796 | (0.0038, 0.158) | 0.049 |
| None vs N95 | 0.312 | (0.240, 0.385) | < 0.0009 |
| Medical mask vs N95 | 0.117 | (0.0618, 0.17) | < 0.0009 |
